# Asthma hospitalisations and heat exposure in England: A case-crossover study during 2002-2019

**DOI:** 10.1101/2022.11.29.22282904

**Authors:** Garyfallos Konstantinoudis, Cosetta Minelli, Holly Ching Yu Lam, Elaine Fuertes, Joan Ballester, Bethan Davies, Ana Maria Vicedo-Cabrera, Antonio Gasparrini, Marta Blangiardo

**Author notes:** **Corresponding author:** Garyfallos Konstantinoudis, MRC Centre for Environment and Health, Department of Epidemiology and Biostatistics, School of Public Health, Imperial College London, London, UK.

## Abstract

**Background:** Previous studies have found an association between warm temperature and asthma hospitalisation. They have reported different sex- and age-related vulnerabilities, nevertheless little is known about how this effect has changed over time and how it varies in space. This study aims to evaluate the association between asthma hospitalisation and warm temperature and investigate vulnerabilities by age, sex, time, and space.

**Methods:** We retrieved individual-level data on summer asthma hospitalisation at high temporal (daily) and spatial (postcodes) resolution during 2002-2019 in England from the NHS Digital. Daily mean temperature at 1km x 1km resolution was retrieved from the UK Met Office. We focused on lags 0-3 days. We employed a case-cross over study design and fitted Bayesian hierarchal Poisson models accounting for possible confounders (rainfall, relative humidity, wind speed, national holidays, and recurrent hospitalisations).

**Results:** After accounting for confounding, we found a 0.85% (95% Credible Interval: 0.64% to 1.07%) increase in the asthma hospitalisation risk for every 1°C increase in the ambient summer temperature. The effect was highest for males aged 15-65 (2.44%, 1.99% to 2.90%). During 2002-2007 we observed a 2.23% (1.86% to 2.60%) increase in hospitalisation risk per 1°C increase in temperature, whereas inconclusive evidence for the periods 2008-2013 and 2014-2019. Populations in Yorkshire and the Humber and East Midlands were the most vulnerable.

**Conclusion:** This study provides evidence of an association between warm temperature and hospital admission for asthma, which was attenuated over time suggesting adaptive mechanisms to heat exposure or differences in lifestyle, comorbid conditions, and occupation over time.

## Introduction

Asthma is the second most prevalent chronic respiratory disease globally (3.6%) and the first in the Western world.^1 2^ The prevalence of asthma in the UK is among the highest in the world with direct NHS costs for managing asthma being estimated at one billion UK pounds per annum with 12% of these costs being for hospital care.^3 4^ Several risk factors, such as smoking, physical activity, medication etc., have been known to trigger asthma.^5^ In addition, environmental risk factors such as inhaling cold air, air-pollutants and allergens can trigger asthma.^5-7^ Previous studies have examined the role of warm temperatures on asthma morbidity, with the results being inconclusive. ^8-16^

The impact of ambient warm temperature on asthma morbidity has received considerable attention recently.^8-16^ Studies in Korea, Shanghai, China, and the Los Angeles County, USA reported weak if any evidence of an association between increasing mean daily or maximum temperature and asthma hospitalisations.^8 10 12^ In contrast, studies in Hong Kong, Himeiji City, Japan, Taiwan, 1816 cities in Brazil, Maryland, USA, New York City, USA and Beijing, China reported an increased asthma hospitalisation risk with higher temperature especially during summer months.^11 13 15-19^

Some of the reasons contributing towards the above-mentioned discrepancies include decisions about the lags of interest, temperature metrics, outcome definitions and selection of confounders, in addition to different adaptation mechanisms (e.g., prevalence of air-conditioning, building infrastructure, etc.), population characteristics (e.g., deprivation, age distribution, etc.) and meteorology across the different countries. Methodological aspects might also limit the generalisability of previous studies. The temporal or geographical resolution in some of the previous studies is coarse leading to a less accurate exposure assignment; most studies examine daily hospitalisation with one study considering monthly hospitalisations, ^14^ and most previous studies assign exposure at the city level ^15-17^. Most studies with individual data were conducted in urban settings, which limits the generalisability of their results given that temperatures in urban settings are typically higher and less variable.^15-17^ One study was nationwide in Taiwan, but the geographical resolution was coarse and the exposure available only at 25 meteorological stations. ^11^ Previous studies have examined effect modification by age and sex, ^15 17^, space ^8 11 16^, and by time, ^16^ but none considered all these dimensions together.

In this nationwide study in England, we examine the effect of ambient temperature on hospital admissions for asthma during 2002-2019. This study aims to address some of the limitations in previous studies by covering one of the largest temporal windows for asthma hospitalisations in the literature, using individual data and exploiting high geographical resolution to link outcome (∼100m) with the exposure (1km). We account for different meteorological confounders and investigate how the effect of ambient temperature on asthma hospitalisations is modified by age and sex. We also examine how the effect is modified by time and region, as we expect the effect to have attenuated over time, due to adaptation (including changes in behaviours, heat-health warnings, etc.), and to be different across space (due to differences in green space, deprivation, etc.).^20 21^

## Methods

### Study population

We retrieved all inpatient hospital admissions for asthma in England during 2002-2019 from NHS Digital Hospital Episode Statistics (HES) data held by the UK Small Area Health Statistics Unit. Age, postcode of residence at time of hospitalisation, and date of hospitalisation were available for each record. We investigated the following main diagnostic group: J45 (Asthma) and J46 (Status asthmaticus) according to the International Classification of Disease version 10 (ICD10).^22^ The analysis is restricted to June, July and August during which the highest temperatures are reached in England.

### Exposure

Daily temperatures at 1km*×*1km resolution were available from the UK Met Office, and obtained with methods described elsewhere.^23^ In brief, the daily temperature in each grid is estimated based on inverse-distance-weighted interpolation of monitoring data, also accounting for latitude and longitude, elevation, coastal influence, and proportion of urban land use. We calculated mean daily temperature taking the average of maximum daily and minimum daily temperatures. To assign daily mean temperature to health records, the postcode centroids of each patient were spatially linked to the 1km*×* 1km grid cell, applying a 100m fuzziness to the postcode location to fulfil governance requirements.^20^ For the main analysis, we averaged the daily mean temperatures across the 3 days prior to the hospital admission (lag0-3), as the delayed effect of heat exposure on health outcomes is expected to last a few days.^15 20 24^

### Confounders

Our main assumptions about possible confounders are shown on the directed acyclic graph (DAG) on Online Supplement Figure S1. Based on the DAG, we decided to account for meteorology (relative humidity, precipitation and wind speed) and national holidays, whereas we assumed that air pollution species (e.g., NO_2_, PM_2.5_, PM_10_ and O_3_) and grass pollen are more likely to mediate, rather than confound, the relationship between temperature and asthma hospitalisation risk.^25^ Mean daily precipitation (*mm*) at 1km*×*1km resolution during 2002 and 2019 was retrieved from the UK Met Office.^23^ Wind speed and relative humidity were retrieved from the UERRA regional reanalysis for Europe and were available daily at 11km x 11km spatial resolution.^26^ National holidays were defined as a binary variable, 0 being a holiday day and 1 otherwise.

### Effect modification

We examined effect modification by age group (0-4, 5-14, 15-64, 65+), sex (males, females), period (2002-2007, 2008-2013 and 2014-2019) and space (nine regions in England, Online Supplemental Figure S2).

### Statistical methods

We used a time-stratified case-crossover design, commonly used for analysing the effect of transient exposures.^27-29^ The temperature on the day of the asthma hospitalisation (event day), is compared with the temperature on non-event days. This design automatically controls for individual level factors that do not vary over time (e.g. age, sex and ethnicity) or vary slowly (e.g. deprivation). We selected non-event days on the same day of week, calendar month and year as the event day to avoid overlap bias,^30^ which also accounts for seasonality (within summer) and long term trends. ^31^

We specified Bayesian hierarchical conditional Poisson models, with a fixed effect on the event/non-event day grouping.^31 32^ The above specification offers an alternative to the conditional logistic model allowing for more flexibility and reducing the computational burden.^32^ We accounted for recurrent hospitalisations using a random effect per patient, specified as a realisation from a normal distribution with zero mean and common variance. To investigate effect modification, we repeated this analysis by age, sex, period, and region.

### Sensitivity analyses

As the effect of temperature on health is typically non-linear,^20 24^ we investigated departures from the linearity assumption. To do so, we modelled temperature using a random walk of order 2, a Gaussian process over the temperature domain with zero mean and covariance that depends on the 2 previous and 2 subsequent observations.^33^ We also examined sensitivity with respect to the lag choice, focusing on any lag between 0 and 5.

All results are reported as medians and 95% Credible Intervals (CrI; 95% probability that the true values lie within this interval) of % increase in hospitalisation risk for every 1°C increase in temperature across the unadjusted and fully adjusted (for rainfall, relative humidity, windspeed and national holidays) models. All analyses are run in R-INLA (Integrated Nested Laplace Approximation).^34^ The code for running the analysis is online available at https://github.com/gkonstantinoudis/asthma_temperature.

## Results

### Population

We retrieved 1,268,725 records with a hospital admission for asthma during 2002-2019 in England. Out of these, 607,089 were readmissions of the same patient. After removing 6,116 duplicate records, 12 records with place of residence outside England and 1,002,512 that did not occur during summer months, we had 260,085 records available for the analysis, Online Supplement Figure S3. We had information on sex for 260,064 admissions, Table 1. Most of the admissions occurred for people between 15 and 65 years old (N = 141,608) and females (N = 147,684), Table 1. We also observed a decreasing trend in hospital admissions for children aged 0-4 with 14,037 and 8,229 reported during the first (2002-2007) and last (2014-2019) period, Table 1, respectively.

**Table 1.**
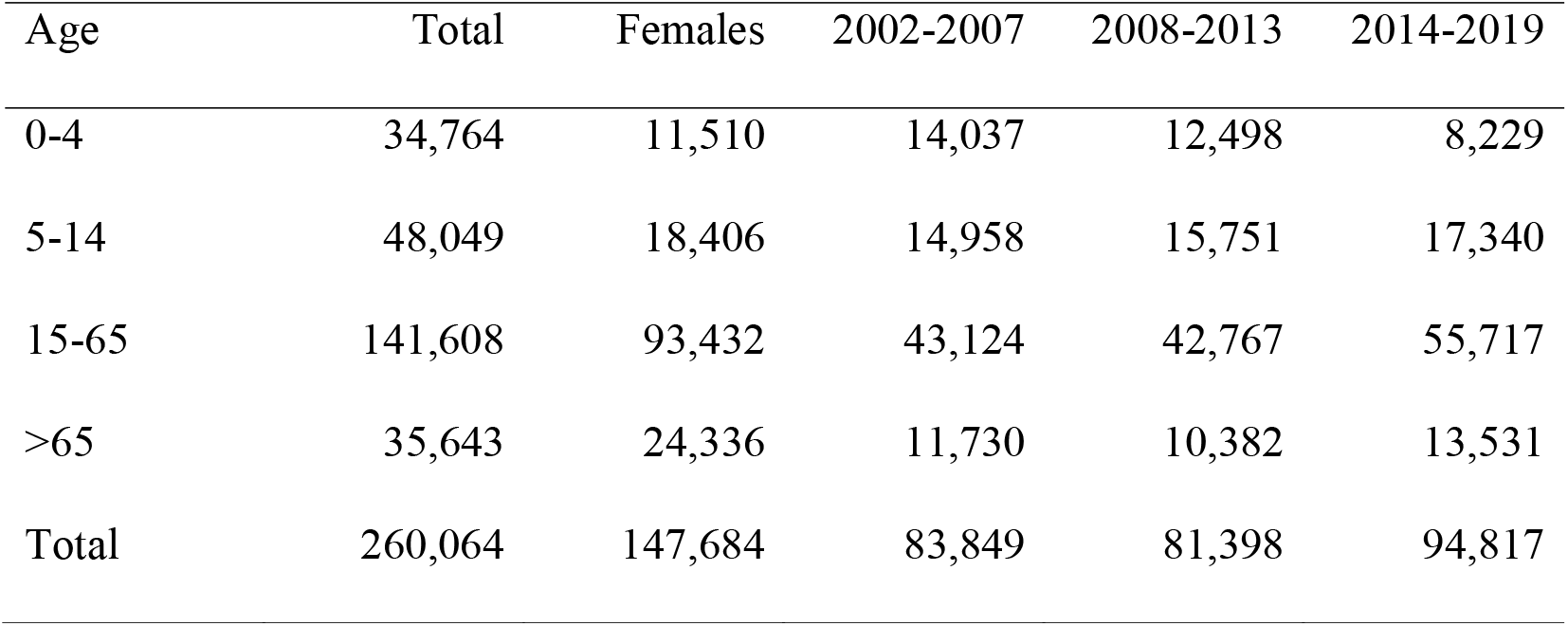
Number of asthma hospitalisation cases by age, sex, and period.

| Age | Total | Females | 2002-2007 | 2008-2013 | 2014-2019 |
| --- | --- | --- | --- | --- | --- |
| 0-4 | 34,764 | 11,510 | 14,037 | 12,498 | 8,229 |
| 5-14 | 48,049 | 18,406 | 14,958 | 15,751 | 17,340 |
| 15-65 | 141,608 | 93,432 | 43,124 | 42,767 | 55,717 |
| >65 | 35,643 | 24,336 | 11,730 | 10,382 | 13,531 |
| Total | 260,064 | 147,684 | 83,849 | 81,398 | 94,817 |

### Exposure

Figure 1 shows the mean of the mean daily temperature across all 1km x 1km grid cells (top panel) and across the summer months by period (2002-2007, 2008-2013 and 2014-2019) in England. The maximum mean daily temperature across England is 24.1°C and was observed in 2019, top panel Figure 1. The mean of the mean daily temperature across England was 16.0°C during 2002-2007, 15.5°C during 2008-2013 and 16.1°C during 2014-2019, red lines top panel Figure 1. The mean of the mean daily temperature across the summer months varied from to 8.9°C to 18.5°C during 2002-2007, from 8.1°C to 17.5°C during 2008-2013 and from 8.0°C to 17.8°C during 2014-2019, bottom panel Figure 1.

**Figure 1.**
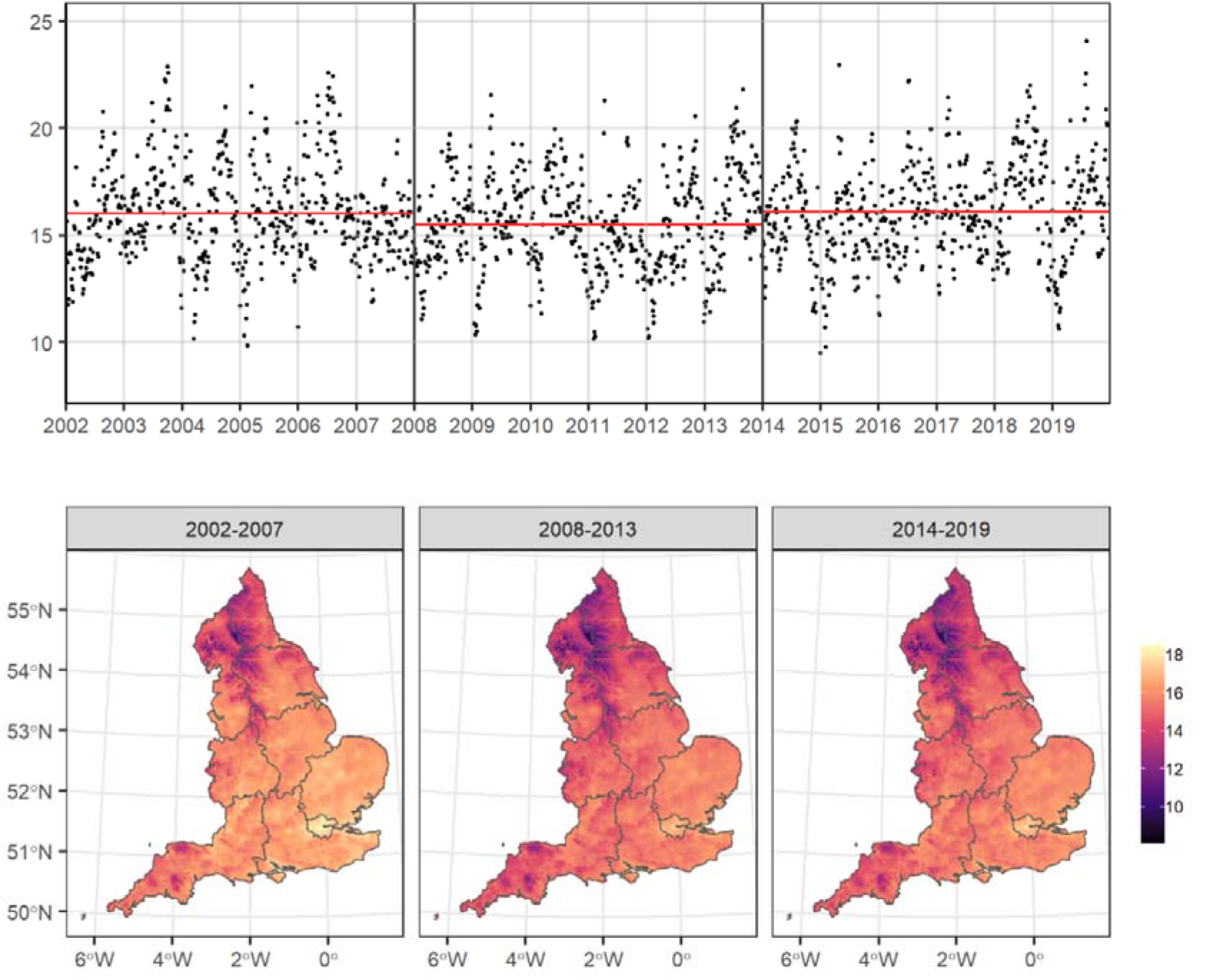
**Top panel:** Mean daily mean temperature (°C) during summer months across England, between 2002 and 2019. The red lines show the mean across the different periods (2002-2007, 2008-2013 and 2014-2019). **Bottom panel:** Mean of daily mean summer temperature (°C) at 1km x 1km grid by period. The grey areas define the regions in England.

### Age and sex effect modification

Figure 2 shows the % change in hospitalisation risk for every 1°C increase in the ambient summer temperature for the different age and sex subgroups during 2002-2019 across the unadjusted and the adjusted models. Accounting for rainfall, relative humidity, windspeed and national holidays does not seem to affect much the observed relationship, Figure 2. Overall, we found a 0.84% (0.65% to 1.04%) and a 0.85% (0.64% to 1.07%) increase in the risk of hospital admission for every 1°C increase in the ambient summer temperature in the unadjusted and adjusted model respectively, Online Supplement Table S1. The effect is consistently higher for males and there is weak, if any, evidence of an effect of temperature for people aged 65 or older. The highest effect was observed for males aged 15-65 with a 1.85% (1.40% to 2.30%) and 2.44% (1.99% to 2.90%) increase in the hospitalisation risk in the unadjusted and adjusted model, Figure 2, and Online Supplement Table S1.

**Figure 2.**
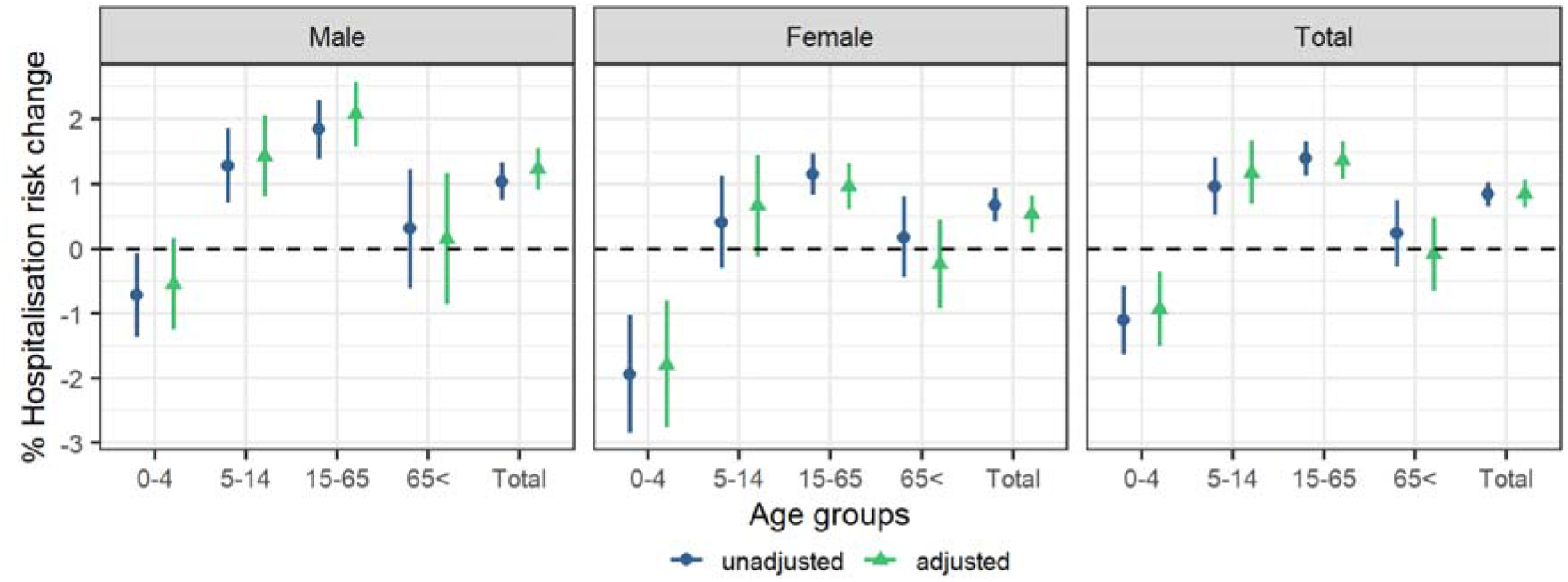
Percentage asthma hospitalisation risk for every 1°C increase in the daily mean summer temperature and 95% credible intervals by sex and age for the unadjusted and fully adjusted (precipitation, relative humidity, wind speed, national holidays, and recurrent hospitalisations) models.

### Temporal effect modification

We examined temporal vulnerabilities of asthma hospitalisation to temperature during summer months. Using the adjusted models, we observe an association of temperature with asthma hospitalisation during 2002-2007, with a 2.23% (1.86% to 2.60%) increase in hospitalisation risk per 1°C increase in temperature, whereas there is inconclusive evidence for the periods 2008-2013 (−0.03%; −0.41% to 0.36%) and 2014-2019 (−0.13%; −0.48 to 0.23%), Figure 3 and Online Supplement Table S2. The total effect in Figure 2 seems to be mainly driven by males, 15-65 years old, during 2002-2007, with a 4.63% (3.77%, 5.49%) increase in the hospitalisation risk for every 1°C increase in temperature, Figure 3 and Online Supplement Table S2. For the rest of the age and sex groups assessed by period the evidence is weaker, Figure 3 and Online Supplement Table S2.

**Figure 3.**
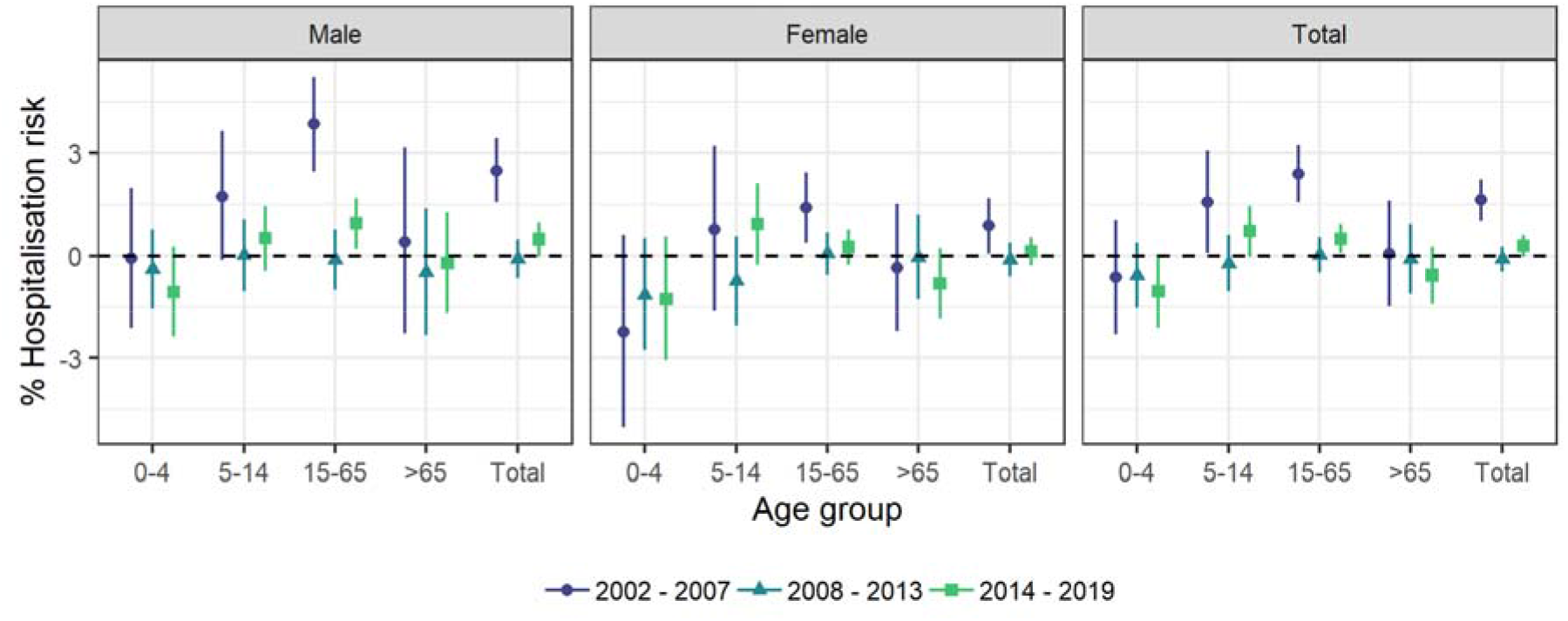
Percentage asthma hospitalisation risk for every 1°C increase in the daily mean summer temperature and 95% credible intervals by sex, age, and period (2002-2007, 2008-2013 and 2014-2019) for the fully adjusted (precipitation, relative humidity, wind speed, national holidays, and recurrent hospitalisations) models.

### Spatial effect modification

Figure 4 presents the spatial vulnerabilities by region and by sex in the adjusted models. We observe that males in Yorkshire and the Humber and East Midlands are the most vulnerable with a % increase in hospitalisation risk being larger than 2% per 1°C increase in temperature in the adjusted models, top panel Figure 4. The 95% CrI of this increase overlaps with that of the main nationwide analysis for this sex group, bottom panel Figure 4. Populations in the South West are consistently the least vulnerable, Figure 4.

**Figure 4.**
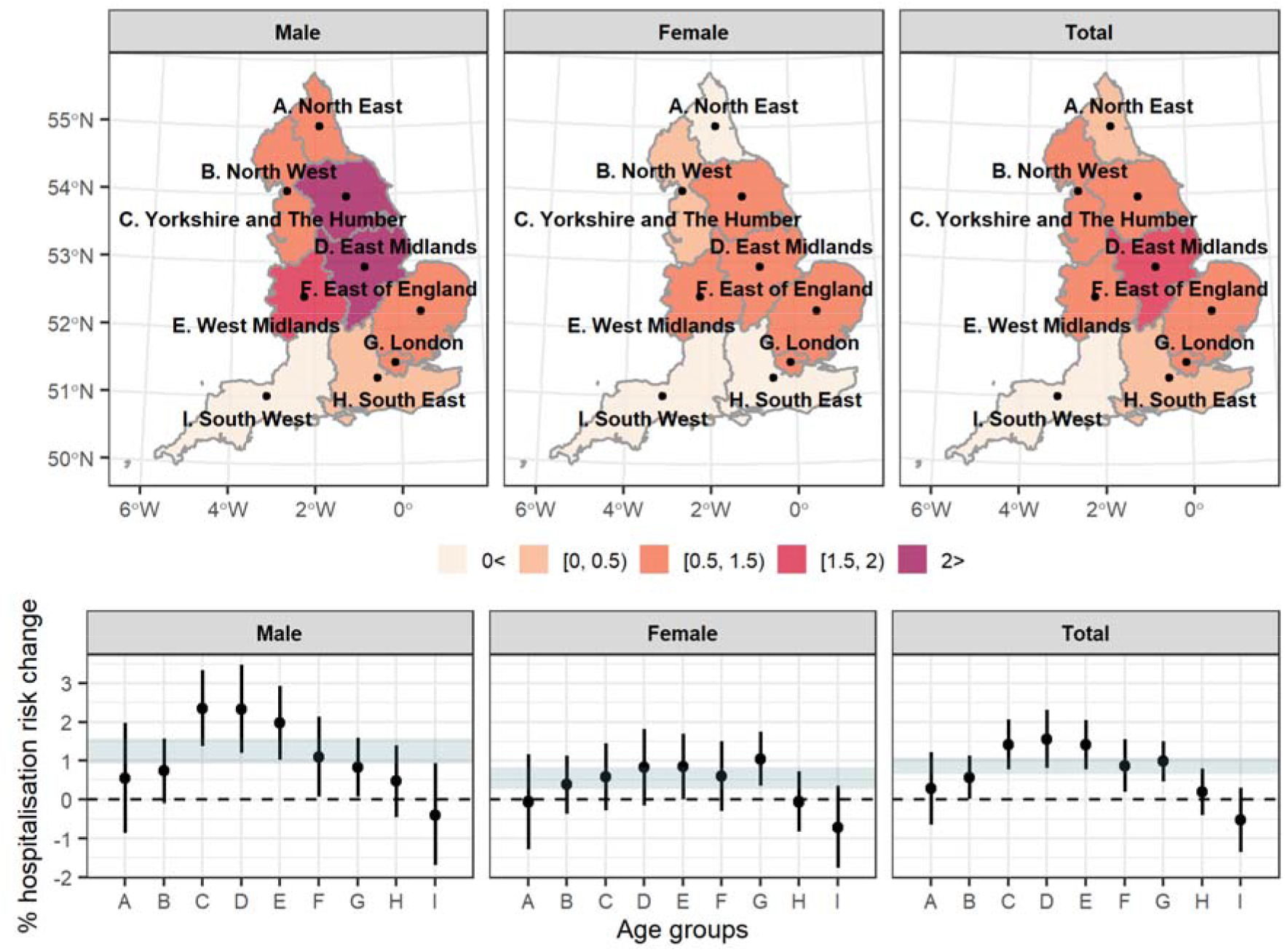
**Top panel:** Percentage asthma hospitalisation risk for every 1°C increase in the daily mean summer temperature by sex and region based on the fully adjusted (precipitation, relative humidity, wind speed, national holidays, and recurrent hospitalisations) models. **Bottom panel:** Percentage asthma hospitalisation risk for every 1°C increase in the daily mean summer temperature and 95% credible intervals (CrI) by region based on the fully adjusted (precipitation, relative humidity, wind speed, national holidays, and recurrent hospitalisations) models. The shaded area illustrates the 95%CrI of the asthma hospitalisation risk nationwide by sex (fully adjusted models).

### Sensitivity analyses

Allowing for flexible fits using random walk of order 2 to model temperature validated our linearity assumption in general, Online Supplement Figure S4. The subgroups deviating from linearity are male between 0 and 4, suggesting weak evidence of a protective effect, and females between 15 and 65, suggesting an effect most prominent for temperatures higher than 20°C, Online Supplement Figure S4. When we examined the different lags we found that the lags contributing most to the effect are 0-3, Online Supplement Figure S5.

### Post-hoc analysis

In a post-hoc analysis we excluded the year 2003 from the 2002-2007 period analysis to examine if the association was driven by the extensive heatwave in 2003. We found that our results were consistent once 2003 was excluded, Online Supplement Figure S6.

## Discussion

In this nationwide study in England investigating the short-term effect of ambient summer temperature on asthma hospitalisations during 2002-2019, we find evidence of an increased risk of asthma hospitalisation associated with increasing temperature. The effect of ambient temperature exposure appears to be modified by sex and age, with males aged between 15 and 65 years being the most vulnerable and children aged between 0 and 4 the most resilient. When we assessed the change in magnitude of this effect over time, we found that the temperature-related asthma hospitalisation risk is largest during 2002-2007, whereas there is weak, if any, effect during 2008-2013 and 2014-2019. We also observed some spatial variation with populations living in the region of Yorkshire and the Humber and East Midlands being most vulnerable.

Our study is comparable with previous studies assessing asthma hospital admissions and ambient temperature during warmer months, rather than studies assessing heat waves and extreme temperatures.^8^ Our study is in line with a time series study in Hong Kong using lag0-3 mean temperature and accounting for relative humidity, wind speed, solar radiation, influenza and air-pollution, which reported an increased asthma hospitalisation relative risk (1.19; 95% Confidence Interval: 1.06, 1.36) at 30°C compared with 27°C during the hot season.^15^ The same study reported that the effect of temperature during the hot season was largest in adults aged between 15 and 59 years.^15^ A time series study examining temperature variation in China and accounting for meteorology reported evidence of an association with childhood asthma admission, but weak evidence for an effect modification by sex.^35^ Another case-crossover study in Detroit reported a protective effect of same day temperature change and emergency department visits for asthma among children.^36^ Our results are in contrast with those of a time series study in Shanghai, China, focusing on different temperature metrics and accounting for meteorology and air-pollution, which reported no association between asthma hospitalisations and warm temperatures.^10^ A case-crossover study in Brazil during 2000-2015 examined temperature variability and asthma hospitalisations and reported a higher effect in people 80 or older, with weak evidence of effect modification by sex.^16^ In our study, weak evidence that people aged 65 or older were vulnerable, while we observed a strong effect modification by sex for temperature-related asthma hospitalisations.

The age modification we observed might have multiple explanations. People aged between 5 and 64 years are likely to be exposed more to environmental triggers in comparison to the older subgroup and children aged less than 5, who stay mostly at home, do not work, and have a less active social life. The adult population could also be more vulnerable due to the higher prevalence of smoking in the adult population resulting in a more severe airway inflammation and lower lung function, which potentially could lead to increased vulnerability to higher temperatures.^15 37^ Although children are expected to be vulnerable to higher temperatures,^38^ we reported a protective effect, which likely reflects a more complex non-linear relationship between temperature and hospital admissions in this age group.

We observed that the effect of ambient temperature on asthma hospitalisations has attenuated over time. The increased risk during 2002-2007 cannot be explained by the extensive heatwave in 2003. This suggests potential adaptive mechanisms to heat exposure over time,^21^ such as more consistent adherence to medication over time, effectiveness of the heat alerts (they were introduced in 2004), infrastructure changes and improved health care. The effect is most prominent in males aged from 15 to 65 years old, which could also point towards sex-related differences in exposure due to differences in behaviours, lifestyle, and occupation, but also could imply differences in the prevalence of other comorbid chronic diseases, such as chronic obstructive pulmonary disease.^39^ Nevertheless, we cannot rule out, potential residual confounding due to data availability or the possibility of this being a chance result, due to the multiple subgroups examined.

In line with our study, previous studies in Taiwan and Brazil have reported effect modification by space.^11 16^ We observed that populations in the regions of Yorkshire and the Humber and East Midlands are the most vulnerable. Potential explanations include spatial effect modifiers that are most prominent in these regions, such as allergen concentration, higher air-pollution levels (particulate matter) and higher prevalence of smoking. Nevertheless, we should iterate that the evidence for heterogeneity in the effect by space is weak as the credible intervals of the effects overlap with the credible intervals of the effect nationwide.

The observed discrepancies with some previous studies can have multiple explanations. The choice of confounding adjustment can impact the results. In addition, the temperature metrics and lags used across the studies are not consistent. Apart from min, mean, max and range of temperature, a couple of studies have used temperature variation.^16 36^ In our study we used the mean, as we were interested in the overall effect of the temperature during the day and were not interested in the impact of night temperatures (using min) or extreme heat during mid-day (using max). It is hard nevertheless to predict how this choice could impact the results, as the different temperature metrics, point towards different potential mechanisms. Previous studies had available coarser geographical resolution (city or county level), which can lead to misclassification of the exposure and inadequate confounding adjustment. In addition, most previous studies are focused on urban settings,^9 12 15-17^ whereas our study provides nationwide estimates. Last, resources for health care setting, health promotion, and infrastructure across the different countries could contribute to the observed discrepancies.

The main strength of our study is the availability of high geographical resolution of the outcome. This allows us to link the outcome with the exposure and confounders with very high precision minimising potential misclassification. As hospitalisations were retrieved from NHS digital (nationwide central data on hospital admissions), we expect to cover most of the asthma hospitalisation burden in England, minimising any selection bias. We used individual-level data, which allows us not only to examine individual-level vulnerabilities, for example related to age and sex, but also to avoid ecological bias.

We assigned temperature exposure using the residential address of the cases and used this as a proxy for individual exposure to temperature. This proxy is expected to be more accurate for children aged 0-4 or adults 65 years or older who are expected to stay longer at home or somewhere in the direct proximity. In addition to this, we used the outdoor temperature which is expected to be different from the temperature inside the house. The misclassification induced by this is not expected to be differential and to lead to bias. Data for relative humidity and wind speed, were only available at coarse geographical resolution, namely at 11km x 11km, which likely misses the localised trends in these covariates and could potentially lead to inadequate adjustment for confounding.

The direct biological mechanism by which higher temperatures trigger asthma exacerbations is unclear. Warm temperatures can aggravate the respiratory system related burden, as they can affect the electrolyte balance.^40^ High temperatures may activate the c-fibre nerve and enhance bronchoconstrictions leading to higher morbidity.^15^ In addition, warmer temperatures can increase allergen or air-pollution (in particular O_3_) concentrations, or help the reproduction and spread of viruses and bacteria causing or aggravating respiratory diseases.^15^

This study provides evidence of an association between warm temperatures and asthma exacerbations in England, and our results are generalisable to developed counties with similar climate, health care and social behaviours. The effect of warm temperatures on asthma hospitalisation has attenuated over time suggesting potential adaptive mechanisms to heat exposure or differences in behaviours, lifestyle, comorbid conditions, other environmental exposures and occupation over time.

## Data Availability

The Small Area Health Statistics Unit (SAHSU) does not have permission to supply data to third parties. For reproducibility purposes we have simulated data and provided the code used for the analysis at https://github.com/gkonstantinoudis/asthma_temperature.

## Acknowledgements

We thank Hima Daby, Gajanan Natu, Eric Johnson and Bethan Davies for their help with data acquisition, storage, preparation, and governance. All authors acknowledge infrastructure support for the Department of Epidemiology and Biostatistics provided by the NIHR Imperial Biomedical Research Centre (BRC). Hospital Episode Statistics data are copyright © 2021, re-used with the permission of NHS Digital. All rights reserved. The Hospital Episode Statistics data were obtained from NHS Digital.

## Contributors

G.K and M.B. conceived the study. M.B. supervised the study. G.K. developed the initial study protocol and discussed it with M.B., C.M., H.C.L. and E.F.. G.K. developed the statistical model, prepared the covariate data, and led the acquisition of hospitalisation data. M.B. validated the code. G.K. ran all the analysis and wrote the initial draft. All the authors contributed in modifying the paper and critically interpreting the results. All authors read and approved the final version for publication. GK is responsible for the overall content as the guarantor.

## Patient consent for publication

Not required.

## Ethics

The analyses were covered by national research ethics approval from the London-South East Research Ethics Committee (Reference 22/LO/0256). Data access was covered by the Health Research Authority Confidentiality Advisory Group under section 251 of the National Health Service Act 2006 and the Health Service (Control of Patient Information) Regulations 2002 (Reference 20/CAG/0028).

