## Supplementary material for "Asthma hospitalisations and heat exposure in England: A case-crossover study during 2002-2019": Online Supplement

### Contents

**S1 Text** **3**

S1.1 Model description . . . . . . . . . . 3

### List of Tables

S1 Percentage hospitalisation risk for every 1$^o$C increase in the temperature and 95% credible intervals by sex and age for the unadjusted and fully adjusted (precipitation and national holidays) models. 4

S2 Percentage hospitalisation risk for every 1$^o$C increase in the temperature and 95% credible intervals by sex, age and period for the fully adjusted (precipitation, relative humidity, wind speed, national holidays and recurrent hospitalisations) model. . . . . . . . . . 5

### List of Figures

S1 Directed acyclic graph for the association between temperature and asthma hospitalisations. . . . 6

S2 Regions in England. . . . . . . . . . 7

S3 Flowchart of the population. . . . . . . . . . 8

S4 Random walks of order 2 on the hospitalisation relative risk by age and sex to allow flexible fits in the unadjusted and adjusted (precipitation, relative humidity, wind speed, national holidays and recurrent hospitalisations) models. The hospitalisation relative risk is relative to the risk at 15$^o$C. 9

S5 Percentage hospitalisation risk for every 1$^o$C increase in the temperature and 95% credible intervals for the fully adjusted (precipitation and national holidays) model across the 0-5 lags. . . . . . . . 10

S6 Percentage hospitalisation risk for every 1$^o$C increase in the temperature and 95% credible intervals for the different age and sex groups during 2002-2007 excluding the 2003 heatwave using the fully adjusted (precipitation and national holidays) model. . . . . . . . . . 11

### S1 Text

#### S1.1 Model description

Let $Y_{mjk}$ be the case-control identifier for the asthma hospitalisation for the event (case or control) at the $m$ grid cell and day, in the $j$-th case-control group and $k$-th patient. Let also $X_m$ be the temperature at $m$ grid cell and day and $Z_m = (1, Z_{1m}, Z_{2m})$ a vector denoting the different confounders. Then:

$$Y_{mjk} \sim \text{Poisson}(\mu_{mjk})$$

$$\log(\mu_{mjk}) = f(X_m) + \alpha Z_m + u_j + w_k$$

$$u_j \sim N(0, 100)$$

$$w_k \sim N(0, \sigma_1^2)$$

In the main analysis we set $f(X_m) = \beta X_m$ whereas for the sensitivity analysis $f(\cdot)$ is the non-linear effect of the $m$ daily temperature in each grid cell. We assume the following second-order random walk (RW2) model:

$$X_{im} \mid X_{(i-1)m}, X_{(i-2)m}, \tau_X \sim \text{Normal}\left(2X_{(i-1)m} + X_{(i-2)m}, \tau_X^{-1}\right), \tag{1}$$

with $\tau_x$ denoting the precision. We fit the above model for the different age and sex groups for the different period and regions.

### Tables

Table S1: Percentage hospitalisation risk for every $1^oC$ increase in the temperature and 95% credible intervals by sex and age for the unadjusted and fully adjusted (precipitation and national holidays) models.

| Age | Sex | Unadjusted | Adjusted |
|---|---|---|---|
| 0-4 | Male | -0.72 (-1.36, -0.07) | -0.54 (-1.24, 0.16) |
| 0-4 | Female | -1.94 (-2.85, -1.03) | -1.79 (-2.77, -0.80) |
| 0-4 | Total | -1.10 (-1.63, -0.58) | -0.92 (-1.50, -0.35) |
| 5-14 | Male | 1.29 ( 0.72, 1.85) | 1.43 ( 0.81, 2.06) |
| 5-14 | Female | 0.41 (-0.30, 1.13) | 0.67 (-0.12, 1.46) |
| 5-14 | Total | 0.97 ( 0.53, 1.41) | 1.18 ( 0.69, 1.67) |
| 15-65 | Male | 1.85 ( 1.40, 2.30) | 2.08 ( 1.58, 2.58) |
| 15-65 | Female | 1.16 ( 0.84, 1.49) | 0.97 ( 0.61, 1.33) |
| 15-65 | Total | 1.40 ( 1.14, 1.67) | 1.37 ( 1.08, 1.66) |
| 65> | Male | 0.32 (-0.61, 1.24) | 0.16 (-0.85, 1.17) |
| 65> | Female | 0.18 (-0.45, 0.81) | -0.24 (-0.92, 0.45) |
| 65> | Total | 0.24 (-0.28, 0.76) | -0.08 (-0.65, 0.49) |
| Total | Male | 1.05 ( 0.75, 1.34) | 1.24 ( 0.91, 1.56) |
| Total | Female | 0.68 ( 0.43, 0.94) | 0.54 ( 0.26, 0.83) |
| Total | Total | 0.84 ( 0.65, 1.04) | 0.85 ( 0.64, 1.07) |

Table S2: Percentage hospitalisation risk for every $1^{o}$C increase in the temperature and 95% credible intervals by sex, age and period for the fully adjusted (precipitation, relative humidity, wind speed, national holidays and recurrent hospitalisations) model.

| Age | Sex | 2002 - 2007 | 2008 - 2013 | 2014 - 2019 |
|---|---|---|---|---|
| 0-4 | Male | -1.09 (-2.17, 0.00) | -0.03 (-1.22, 1.17) | -1.25 (-2.68, 0.19) |
| 0-4 | Female | -3.35 (-4.87, -1.81) | -0.72 (-2.40, 0.98) | -2.10 (-4.02, -0.16) |
| 0-4 | Total | -1.76 (-2.64, -0.86) | -0.15 (-1.13, 0.84) | -1.46 (-2.62, -0.29) |
| 5-14 | Male | 2.93 ( 1.86, 4.02) | 0.61 (-0.49, 1.72) | -0.41 (-1.46, 0.65) |
| 5-14 | Female | 0.87 (-0.54, 2.29) | -0.14 (-1.51, 1.25) | 0.39 (-0.91, 1.71) |
| 5-14 | Total | 2.32 ( 1.46, 3.18) | 0.41 (-0.45, 1.29) | -0.03 (-0.86, 0.80) |
| 15-65 | Male | 4.63 ( 3.77, 5.49) | -0.26 (-1.17, 0.66) | 0.61 (-0.21, 1.45) |
| 15-65 | Female | 2.93 ( 2.28, 3.58) | -0.14 (-0.78, 0.51) | -0.07 (-0.64, 0.51) |
| 15-65 | Total | 3.63 ( 3.11, 4.15) | -0.15 (-0.68, 0.38) | 0.19 (-0.29, 0.66) |
| 65> | Male | 0.54 (-1.13, 2.23) | -0.65 (-2.54, 1.27) | -0.34 (-1.98, 1.32) |
| 65> | Female | 1.13 (-0.03, 2.32) | -0.58 (-1.83, 0.69) | -1.48 (-2.61, -0.33) |
| 65> | Total | 1.05 ( 0.08, 2.02) | -0.50 (-1.55, 0.57) | -1.03 (-1.97, -0.08) |
| Total | Male | 2.62 ( 2.08, 3.17) | 0.12 (-0.46, 0.71) | 0.07 (-0.50, 0.63) |
| Total | Female | 1.84 ( 1.34, 2.35) | -0.18 (-0.68, 0.33) | -0.28 (-0.75, 0.18) |
| Total | Total | 2.23 ( 1.86, 2.60) | -0.03 (-0.41, 0.36) | -0.13 (-0.48, 0.23) |

### Figures

Figure S1: Directed acyclic graph for the association between temperature and asthma hospitalisations.

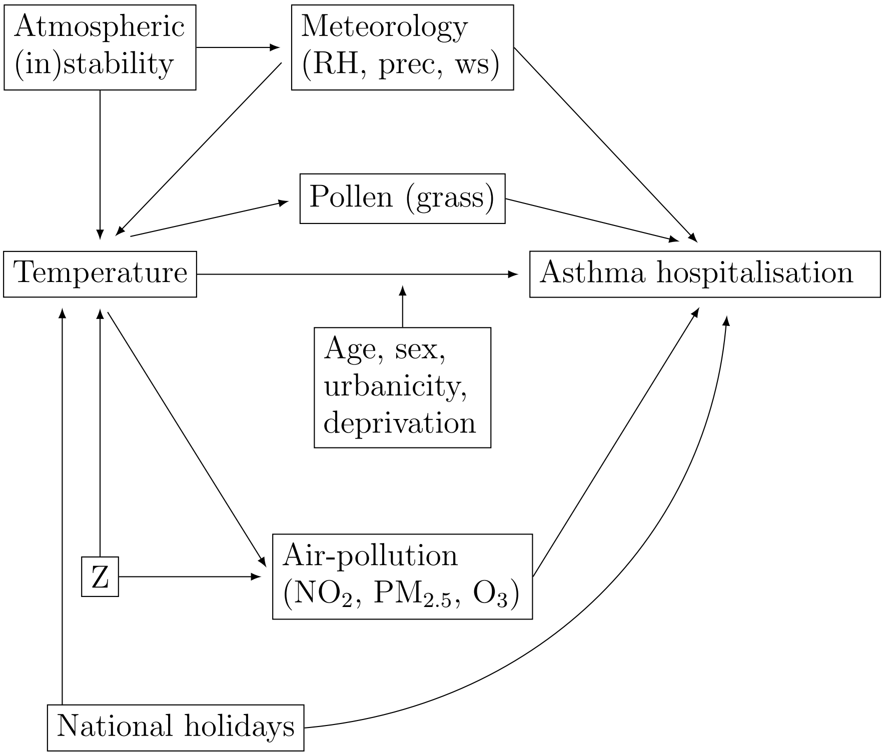


* Abbreviations: prec, precipitation; ws, wind speed,

Figure S2: Regions in England.

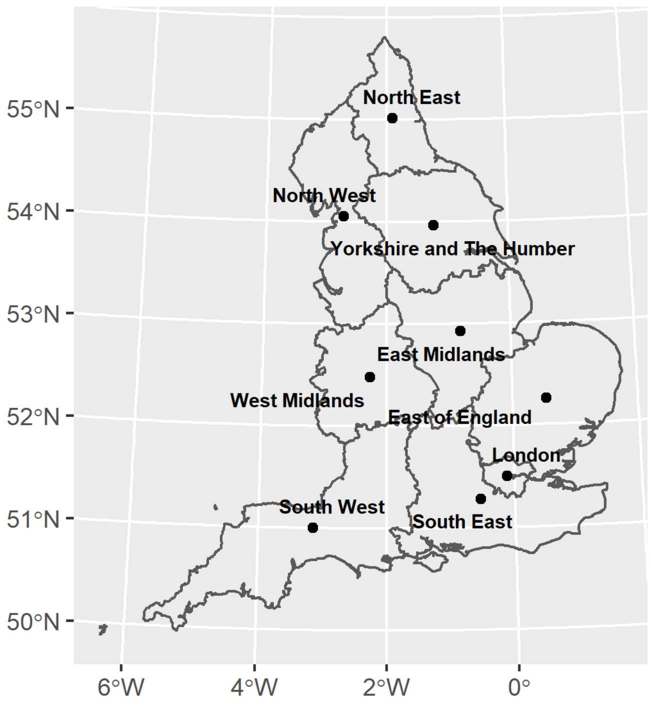

Figure S3: Flowchart of the population.

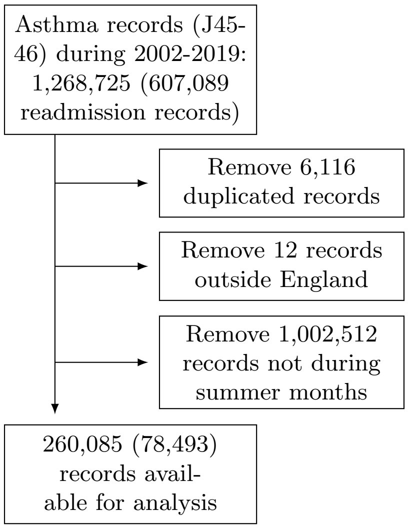

Figure S4: Random walks of order 2 on the hospitalisation relative risk by age and sex to allow flexible fits in the unadjusted and adjusted (precipitation, relative humidity, wind speed, national holidays and recurrent hospitalisations) models. The hospitalisation relative risk is relative to the risk at 15$^{o}$C.

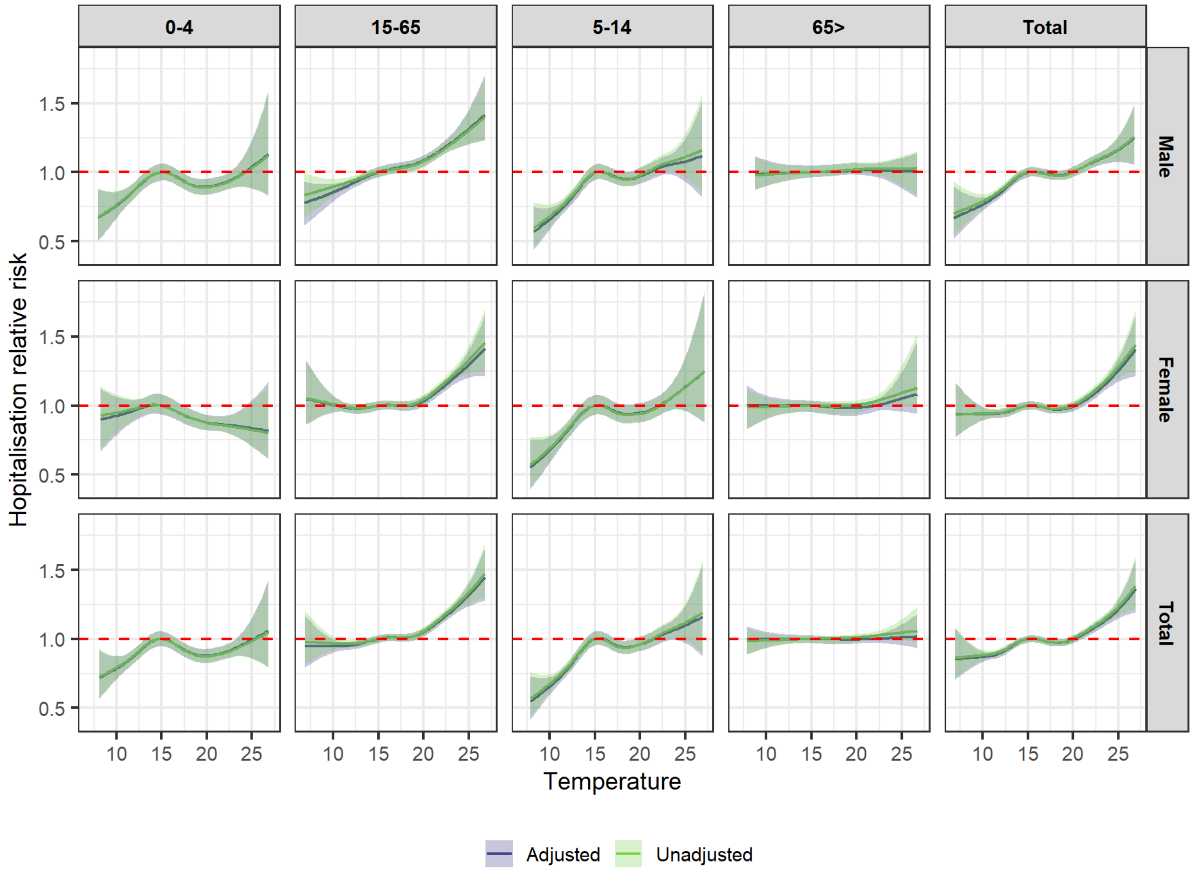

Figure S5: Percentage hospitalisation risk for every 1$^o$C increase in the temperature and 95% credible intervals for the fully adjusted (precipitation and national holidays) model across the 0-5 lags.

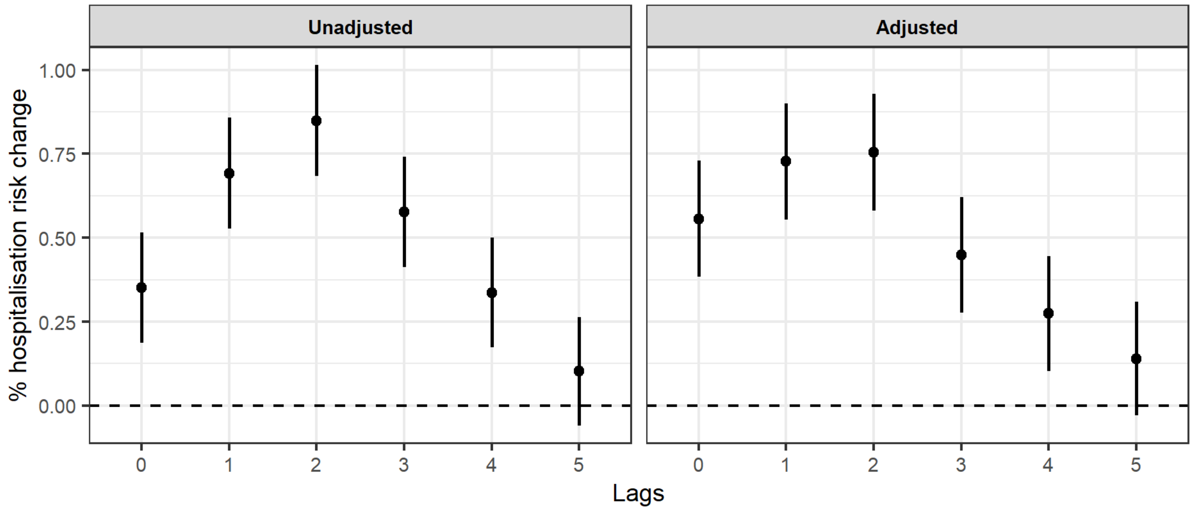

Figure S6: Percentage hospitalisation risk for every 1$^o$C increase in the temperature and 95% credible intervals for the different age and sex groups during 2002-2007 excluding the 2003 heatwave using the fully adjusted (precipitation and national holidays) model.

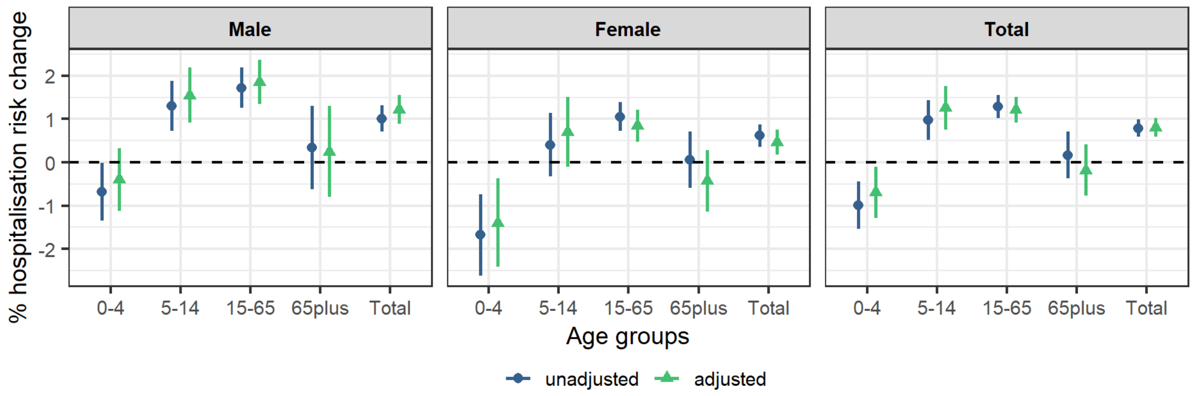
